# Examining the impact of outdoor walk group attendance on health among older adults with mobility limitations in the Getting Older Adults Outdoors (GO-OUT) randomized trial

**DOI:** 10.1101/2024.08.22.24312456

**Authors:** Tai-Te Su, Ruth Barclay, Rahim Moineddin, Nancy M. Salbach

## Abstract

**Objective:** The Getting Older Adults Outdoors randomized trial showed a 10-week outdoor walk group (OWG) program was not superior to 10 weekly phone reminders on increasing physical and mental health; however, OWG attendance varied. This study examined whether a dose-response relationship existed between OWG attendance and improvement in physical and mental health among older adults with mobility limitations.

**Methods:** We analyzed data from 98 older adults randomized to a 10-week park-based OWG program. Participants were classified as attending 0–9, 10–15, and 16–20 OWG sessions based on attendance tertiles. Outcomes included change in scores on measures of walking endurance, comfortable and fast walking speed, balance, lower extremity strength, walking self-efficacy, and emotional well-being pre- to post-intervention.

**Results:** Seventy-nine older adults with complete information on the seven health outcomes were included (age=74.7±6.6 years, 72% female). Compared to those who attended 0–9 OWG sessions, participants attending 16–20 sessions exhibited a 52.7-meter greater improvement in walking endurance (95% CI:12.3, 93.1); 0.15-meter/second greater improvement in comfortable walking speed (95% CI:0.00, 0.29); and 0.17-meter/second greater improvement in fast walking speed (95% CI:0.02, 0.33). Higher attendance was also associated with higher odds of experiencing an improvement in walking self-efficacy (OR=4.03; 95% CI:1.05, 16.85) and fast walking speed (OR=9.00, 95% CI:1.59, 61.73). No significant dose-response relationships for balance, lower extremity strength, and emotional well-being were observed.

**Conclusions:** Higher attendance in outdoor walking interventions is associated with greater improvements in walking endurance, walking speed, and walking self-efficacy among older adults with mobility limitations.

## INTRODUCTION

Walking in outdoor natural environments plays a critical role in supporting the health and well-being of older adults. An accumulating body of evidence has shown that outdoor walking is associated with a wide array of health benefits, including improved physical activity and fitness [1,2] better cognitive and mental functioning [3,4], along with increased energy and greater feelings of revitalization [5]. In addition, exposure to natural environments is associated with a decreased incidence of chronic health conditions such as diabetes, stroke, coronary heart disease, and all-cause mortality [6,7]. The combination of physical activity and natural environments makes outdoor walking an ideal and promising strategy to promote healthy aging.

Although walking outdoors is a preferred mode of physical activity [5], a recent systematic review and meta-analysis found no significant difference between outdoor community ambulation and other comparison interventions (e.g., standard care or educational lectures) in improving older adults’ walking endurance and depression [8]. While the limited number of studies included and quality of evidence partly account for this phenomenon [8], attendance may play a major role in shaping the overall effects of outdoor walking. In particular, previous studies have shown that attendance, defined as the number or proportion of sessions attended, can fluctuate between 58% and 77% among older adults participating in exercise interventions [9–14]. Given that attendance levels can affect the total amount of training and environmental stimuli received, it is important to understand whether and how the effects of outdoor walking may vary depending on participants’ attendance to the interventions.

The Getting Older Adults Outdoors (GO-OUT) study [15] provides a basis for examining the impact of outdoor walking attendance on older adults’ health. The GO-OUT trial is a two-group randomized controlled trial designed to evaluate the effectiveness of a park-based, task-oriented walking program on walking activity and capacity among older adults with mobility limitations [15]. One group participated in a 1-day educational workshop plus a 10-week outdoor walk group (OWG) program, while the other group received the same workshop plus 10 weekly telephone reminders. The trial was conducted across four large Canadian cities, and outcome assessments were performed at baseline, 3 months, 5.5 months, and 12 months. Despite participants providing positive feedback during the interviews [16], our quantitative analyses revealed no significant difference between the two groups in changes in minutes walked outdoors pre-to post-intervention [17]. Further, the impact of the OWG was also not superior to the weekly reminders in improving indicators of walking capacity, health-promoting behaviors, and successful aging with one exception. The OWG led to greater improvement in walking self-efficacy compared to the weekly reminders group [17]. As noted in the process evaluation of the GO-OUT trial [18], the observed lack of effects may be attributed to varying levels of attendance within the OWG, as mean attendance ranged from 43.8% to 84.9% across the study sites.

To better understand the benefits of outdoor walking interventions and inform their implementation in community settings, the objective of this study was to explore whether a dose-response relationship existed between attendance in the OWG program and improvements in health among older adults with mobility limitations. We focused on seven physical and mental health outcomes measured in the GO-OUT trial: walking endurance, comfortable and fast walking speed, balance, lower extremity strength, walking self-efficacy, and emotional well-being. These outcomes were investigated because of their relevance to the core components of the OWG program and their reciprocal relationship with outdoor walking behavior among older adults [16,19,20]. Knowledge derived from the present study will not only help to equip clinical practitioners with evidence-based recommendations but also inform marketing strategies to motivate older adults to engage with community-based exercise programs.

## METHODS

### Study Design and Participants

The GO-OUT randomized controlled trial was conducted across four urban cities in Canada: Edmonton, Winnipeg, Toronto and Montreal, and was registered on ClinicalTrials.gov (registration number: NCT03292510). Between February 20, 2018 and June 15, 2019, the GO-OUT trial enrolled 190 older adults aged 65 years and older, living independently in the community, who self-reported difficulty walking outdoors but affirmed the capacity to walk at least one block (50m) with or without a walking aid, expressed willingness to sign a liability waiver or obtain physician clearance for exercise, exhibited mental competency (scoring at least 18 out of 22 on the Mini-Mental State Exam telephone version), and were able to speak and understand English [15].

Upon enrollment, participants attended a 1-day educational workshop designed to enhance knowledge and skills to engage in outdoor walking and prevent falls. After the workshop, participants were stratified by study site and participant type (enrolled as an individual or a dyad) and randomized into either the 10-week weekly reminders group (n=92) or the 10-week OWG program (n=98). Outcome assessments were performed at baseline, 3 months, 5.5 months, and 12 months. As this study primarily focused on the dose-response relationship between attendance in the OWG and improvement in health, we utilized attendance data and limited our analytic sample to participants randomly assigned to the OWG program with complete data in all seven health outcomes at month 3 (n=79). Research ethics boards at each site approved the trial protocol and participants provided written informed consent prior to baseline assessment. Detailed information on the GO-OUT trial was described in the study protocol [15].

### Outdoor Walk Group Program and Attendance

Building upon a conceptual framework of community mobility [21], the OWG program was designed to enhance older adults’ competency in multiple dimensions of mobility such as walking distance, walking speed, postural transitions, and external physical load, within an outdoor environment [15]. The program involved two 1-hour sessions of group-based outdoor walking in large parks each week for a duration of 10 weeks (maximum of 20 sessions). The OWG program was progressive, task-specific, and implemented during summer months (June– August). Each session consisted of a 10-minute warm up, a distance walk, practice of a specific outdoor walking skill, a second distance walk, and a 10-minute cool down. Each OWG had a leader (physiotherapist or kinesiologist), who was authorized to adjust the program’s difficulty to ensure an optimal level of challenge for the participants [18]. Leaders were supported by additional trained staff to ensure a 1:3 ratio of OWG facilitators (leader/assistant) to participants.

After each OWG session, group leaders completed standardized forms to document implementation of session activities and participant attendance [18]. For the analysis of this study, participants were grouped into three categories based on attendance tertiles, which reflected the tri-modal distribution of attendance in the OWG program. The first, second, and third tertiles corresponded to participants attending 0–9, 10–15, and 16–20 total OWG sessions, respectively.

### Outcomes for Dose-Response Analysis

### Walking Endurance

Walking endurance was assessed using the 6-minute walk test [22]. Participants received instructions to walk as far as possible in six minutes by walking back and forth along a straight, 30-meter walkway. Participants were asked to complete the test using their assistive devices and corrective eyewear as applicable. The maximum distance walked within six minutes was documented in meters. The 6-minute walk test demonstrates excellent test-retest reliability (intra-class correlation coefficients ICC=0.95) and is considered a valid test of physical functioning and endurance among older adults [23,24].

### Walking Speed at Comfortable and Fast Paces

Walking speed was assessed using the 10-meter walk test [25]. Participants were instructed to walk over a 14-meter walkway twice, once at a comfortable and once at a fast pace. Participants were asked to use their usual assistive devices and corrective eyewear to complete the test as applicable. The time taken to walk the central 10 meters was documented in seconds and used to calculate comfortable and fast walking speed (meter/second). The 10-meter walk test demonstrates excellent test-test reliability (ICCs=0.96-0.98) and is recommended for clinical assessment of walking speed among older adults [26,27].

### Balance

Balance was evaluated using the Mini Balance Evaluation System test (mini-BESTest) [28]. The mini-BESTest is a 14-item test developed to assess four balance domains [29]. Scores for anticipatory postural adjustments, reactive postural control, and sensory orientation range from 0 to 6, whereas the score for dynamic gait ranges from 0 to 10. Higher scores indicate greater balance in the respective subsystem. A total summed score, ranging from 0 to 28, is calculated to measure overall balance function. The mini-BESTest demonstrates good to excellent reliability (ICC>0.90). Evidence of construct validity has been reported [28,30].

### Lower Extremity Strength

The 30-second sit-to-stand test [31] was adopted as a proxy measure of lower extremity muscle strength. Participants were instructed to sit in the middle of the chair, place arms folded across the chest, keep their feet placed on the floor, and repeat rising to a full standing position and sitting back down. The number of sit-to-stands completed in 30 seconds was documented. The 30-second sit-to-stand test has shown excellent test-retest reliability (ICC=0.95) and validity among community-dwelling older adults [31,32].

### Walking Self-Efficacy

The ambulatory self-confidence questionnaire [33] was used to evaluate walking self-efficacy. A total of 22 items were asked to measure how confident participants are in their ability to walk in different environmental situations. Participants rated their responses to each item on a 10-point scale (0 not at all confident to 10 extremely confident). The total score is calculated as the mean of item scores and ranges from 0 to 10, with higher scores indicating greater confidence with walking ability. The ambulatory self-confidence questionnaire demonstrates excellent internal consistency (Cronbach’s α=0.95) and test-retest reliability (ICC=0.92) among community-dwelling older adults [33].

### Emotional Well-Being

We used the emotional well-being scale within the RAND 36-Item Health Survey (RAND-36) to measure participants’ emotional well-being [34]. Participants answered five questions on their emotions and experiences over the past four weeks. Total scores range from 0 to 100, with higher scores indicating better emotional well-being. The RAND-36 Health Survey was adapted from the instruments administered in the Medical Outcome Study [34], and its emotional well-being scale has demonstrated internal consistency and reliability (Cronbach’s α=0.90).

### Participant Characteristics

Information on individual characteristics was collected at baseline [15]. Participants self-reported their age (years), sex (male vs. female), highest level of education, and use of walking aids. The Charlson Comorbidity Index [35] was used to assess participants’ diagnosis of 18 comorbidities such as diabetes, hypertension, cancer, and heart diseases at baseline. The final weighted score ranges from 0 to 39, with higher scores indicating the presence of greater and more severe comorbidities.

### Statistical Analysis

Descriptive statistics were computed to describe participant characteristics and measures of physical and mental health at baseline in each attendance tertile groups (i.e., attended 0–9, 10– 15, and 16–20 sessions). We conducted nonparametric Kruskal-Wallis and chi-squared tests to compare differences in baseline characteristics and health between the tertile groups.

To examine potential dose-response relationships between attendance and improvement in health among older adults enrolled in the OWG program, we followed established methodologies and operationalized the definition of response using three approaches: the extent of improvement [36], the odds of improvement [37], and the overall count of improvement in health [38]. *Extent of improvement*: First, we calculated absolute change scores on each of the seven health outcomes (walking endurance, comfortable and fast walking speed, balance, lower extremity strength, walking self-efficacy, and emotional well-being) from baseline to 3 months, where a positive change score indicates an improvement in the respective outcome. We employed linear regression models, using change scores as the dependent variable, and regressed them against the three attendance tertile groups. This allowed us to investigate whether higher attendance was associated with a greater extent of improvement in physical and mental health. A total of seven regression models were tested, with each focusing on a specific health outcome. In addition, we reported Hedges’ *g* effect sizes to provide insights into the magnitude of the dose-response relationship tested, with values of 0.15, 0.40, 0.75 representing small, medium, and large effects [39]. *Odds of improvement*: Second, we created a binary response variable to represent improvement (yes for change score > 0; no for change score ≤ 0) in each of the seven health outcomes. Separate logistic regression models were implemented to compare the odds of experiencing improvement in each health outcome across the three attendance groups. *Overall count of improvement in health*: Third, an overall count of improved health outcomes was created for each participant. A score of 0 suggests no improvement was found, while a score of 7 indicates improvement in all seven physical and mental health outcomes. A Poisson regression model was used to assess whether the number of improved health outcomes increased as a function of attendance in the OWG program. All models were adjusted for participant sex and study site, which were the two characteristics that differed across the attendance groups.

Recognizing that our analytic sample was restricted to 79 out of 98 OWG participants who had complete data on all seven health outcomes at month 3, we employed the two-stage Heckman correction [40,41] to account for potential selection bias. Specifically, we first computed the inverse Mills ratio based on the full sample of older adults who were assigned to the outdoor walking group (n=98) using a probit model. This ratio was then included in the regression models as an explanatory variable to adjust for non-selection hazards [40]. We conducted sensitivity analysis to compare findings with and without the Heckman correction. A two-tailed p-value less than 0.05 was considered statistically significant. All statistical analyses were performed using R software (Version 4.0.5).

## RESULTS

### Descriptive Statistics

Table 1 presents baseline participant characteristics and health outcomes for the entire sample and by level of outdoor walk group attendance of the 79 older adults included in the dose-response analysis. Mean age was 74.7 years (standard deviation [SD]=6.6), and 72% of the participants were female. Prior to the OWG program, the mean distance that participants achieved on the 6-minute walk test was 362.6 meters (SD=91.8). The average walking speed was 1.09 m/s (SD=0.24) at a comfortable pace and 1.43 m/s (SD=0.31) at a fast pace. The mean score of mini-BESTest, 30-second sit-to-stand test, and ambulatory self-confidence questionnaire was 20.6 (SD=4.7; out of 28), 8.2 (SD=3.5), and 7.9 (SD=1.6; out of 10), respectively. Upon closer examination of the three attendance tertile groups, participants from the Montreal (53%) and Winnipeg (56%) sites attended 16 or more outdoor walking sessions more frequently than those from Edmonton (16%) and Toronto (15%) sites (*p*=0.007). Additionally, around 55% of the 28 male participants but only 28% of the 72 female participants attended 16 or more outdoor walking sessions (*p*=0.02). Remaining baseline individual and clinical characteristics did not differ across the three attendance tertile groups.

**Table 1.** Participant characteristics and performance on health outcome measures at baseline by level of outdoor walk group attendance.

| Participant characteristics<br>(units or scoring) | Pooled<br>(n = 79) | OWG attendance tertile* |  |  | p value |
| --- | --- | --- | --- | --- | --- |
|  |  | 1<sup>st</sup> tertile (n = 17) | 2<sup>nd</sup> tertile (n = 34) | 3<sup>rd</sup> tertile (n = 28) |  |
|  |  | Mean ± SD / n (%) |  |  |  |
| Study sites |  |  |  |  | .007 |
| Edmonton | 19 (24) | 8 (42) | 8 (42) | 3 (16) |  |
| Montreal | 15 (19) | 1 (7) | 6 (40) | 8 (53) |  |
| Toronto | 20 (25) | 6 (30) | 11 (55) | 3 (15) |  |
| Winnipeg | 25 (32) | 2 (8) | 9 (36) | 14 (56) |  |
| Participant type |  |  |  |  | .45 |
| Individual | 61 (77) | 15 (25) | 24 (39) | 22 (36) |  |
| Dyad | 18 (23) | 2 (11) | 10 (56) | 6 (33) |  |
| Cohort |  |  |  |  | .81 |
| 2018-19 | 29 (37) | 7 (24) | 13 (45) | 9 (31) |  |
| 2019-20 | 50 (63) | 10 (20) | 21 (42) | 19 (38) |  |
| Age years) | 74.7 ± 6.6 | 73.5 ± 6.5 | 74.4 ± 6.3 | 75.8 ± 7.1 | .58 |
| Sex |  |  |  |  | .02 |
| Male | 22 (28) | 1 (5) | 9 (41) | 12 (55) |  |
| Female | 57 (72) | 16 (28) | 25 (44) | 16 (28) |  |
| Educational attainment |  |  |  |  | .37 |
| Secondary or lower | 19 (24) | 3 (16) | 6 (32) | 10 (53) |  |
| Some college | 29 (37) | 5 (17) | 14 (48) | 10 (34) |  |
| Bachelor's degree or higher | 31 (39) | 9 (29) | 14 (45) | 8 (26) |  |
| Uses a walking aid | 22 (28) | 5 (23) | 11 (50) | 6 (27) | .63 |
| Charlson comorbidity index (0–39) | 2.1 ± 2.0 | 2.1 ± 3.0 | 2.2 ± 1.6 | 1.9 ± 1.7 | .40 |
| 6-minute walk test (m) | 362.6 ± 91.8 | 401.7 ± 87.5 | 346.0 ± 91.7 | 360.1 ± 90.8 | .14 |
| 10-meter walk test at comfortable pace (m/s) | 1.09 ± 0.24 | 1.21 ± 0.26 | 1.06 ± 0.22 | 1.04 ± 0.24 | .07 |
| 10-meter walk test at fast pace (m/s) | 1.43 ± 0.31 | 1.56 ± 0.32 | 1.37 ± 0.28 | 1.41 ± 0.34 | .12 |
| Mini-BESTest (0–28) | 20.6 ± 4.7 | 21.4 ± 4.4 | 20.2 ± 4.6 | 20.6 ± 5.0 | .71 |
| 30-second sit-to-stand (# stands) | 8.2 ± 3.5 | 8.3 ± 3.8 | 8.1 ± 3.5 | 8.1 ± 3.6 | .98 |
| ASCQ (0–10) | 7.9 ± 1.6 | 8.1 ± 1.6 | 7.6 ± 1.7 | 8.2 ± 1.4 | .36 |
| RAND-36 emotional well-being (0–100) | 76.4 ± 16.9 | 69.7 ± 20.8 | 79.7 ± 13.6 | 76.6 ± 17.3 | .28 |

*Note:* OWG = outdoor walk group; Mini-BESTest = Mini Balance Evaluation System test; ASCQ = ambulatory self-confidence questionnaire; SD = standard deviation; m = meters; m/s = meters/second. \*The 1<sup>st</sup> tertile group attended 0–9 sessions; the 2<sup>nd</sup> tertile group attended 10–15 sessions; the 3<sup>rd</sup> tertile group attended 16–20 sessions. Tertiles were calculated based on all enrolled participants in the outdoor walk group program (n=98). Values in the parenthesis indicate row percentages.

### Influence of Outdoor Walk Group Attendance on the Extent of Improvement in Health

The associations between attendance in the OWG program and extent of improvement in the seven physical and mental health outcomes are summarized in Table 2. Compared to those who attended 0–9 outdoor walking sessions (1^st^ tertile), participants who attended 16–20 sessions (3^rd^ tertile) exhibited, on average, a 52.72-meter greater improvement in the 6-minute walk test performance from baseline to 3 months (95% CI: 12.31, 93.13, *p*=0.01). A similar yet nonsignificant dose-response pattern was observed, where participants attending 16–20 sessions exhibited a tendency towards greater improvement in walking endurance than those who attended 10–15 sessions (*b*=27.28, 95% CI: -2.10, 56.66, *p*=0.07). The effect sizes of the dose-response relationships observed were small in magnitude (Hedges’ *g* range: 0.28–0.36; see Supplementary Table 1).

**Table 2.** Associations between outdoor walk group attendance and the extent of improvement in health outcome measures from baseline to 3 months.

| Health outcome measures<br>(units or scoring) | Comparisons between OWG attendance tertiles |  |  |
| --- | --- | --- | --- |
|  | 2<sup>nd</sup> tertile (10–15 sessions) | 3<sup>rd</sup> tertile (16–20 sessions) | 3<sup>rd</sup> tertile (16–20 sessions) |
|  | vs | vs | vs |
|  | 1<sup>st</sup> tertile (0–9 sessions) | 1<sup>st</sup> tertile (0–9 sessions) | 2<sup>nd</sup> tertile (10–15 sessions) |
|  | Unstandardized regression coefficient *b* [95% confidence intervals] |  |  |
| 6-minute walk test (m) | 25.44 [-10.07, 60.94] | 52.72 [12.31, 93.13] * | 27.28 [-2.10, 56.66] † |
| 10-meter walk test at comfortable pace (m/s) | 0.10 [-0.03, 0.22] | 0.15 [0.00, 0.29] * | 0.05 [-0.05, 0.15] |
| 10-meter walk test at fast pace (m/s) | 0.14 [0.00, 0.27] * | 0.17 [0.02, 0.33] * | 0.04 [-0.07, 0.15] |
| Mini-BESTest (0–28) | -1.10 [-3.37, 1.16] | 0.02 [-2.59, 2.63] | 1.13 [-0.74, 2.99] |
| 30-second sit-to-stand (# stands) | -0.33 [-1.88, 1.22] | 0.29 [-1.50, 2.07] | 0.62 [-0.66, 1.89] |
| ASCQ (0–10) | 0.39 [-0.40, 1.18] | 0.48 [-0.46, 1.41] | 0.09 [-0.61, 0.78] |
| RAND-36 emotional well-being (0–100) | -6.70 [-16.31, 2.91] | -2.79 [-14.03, 8.46] | 3.91 [-4.46, 12.29] |

*Note:* The regression coefficients are in reference to the lower tertile/attendance group, with positive values suggesting potential dose-response relationships. All regression models were adjusted for participant sex, study site, and the Heckman correction. †*p*<0.10; \**p*<0.05

Participants attending 10–15 outdoor walking sessions exhibited a greater extent of improvement in fast walking speed compared to those with 0–9 sessions (*b*=0.14, 95% CI: 0.00, 0.27, *p*=0.048). Similarly, those attending 16–20 sessions demonstrated greater improvements in both comfortable walking speed (*b*=0.15, 95% CI: 0.00, 0.29, *p*=0.04) and fast walking speed (*b*=0.17, 95% CI: 0.02, 0.33, *p*=0.03) compared to participants with 0–9 sessions. The effects observed were medium to strong in magnitude (Hedges’ *g* range: 0.48–0.80). There were no significant relationships found between attendance and absolute changes in measures of balance, lower extremity strength, walking self-efficacy, and emotional well-being. Fig 1 presents results without covariate adjustment.

**Fig 1.**
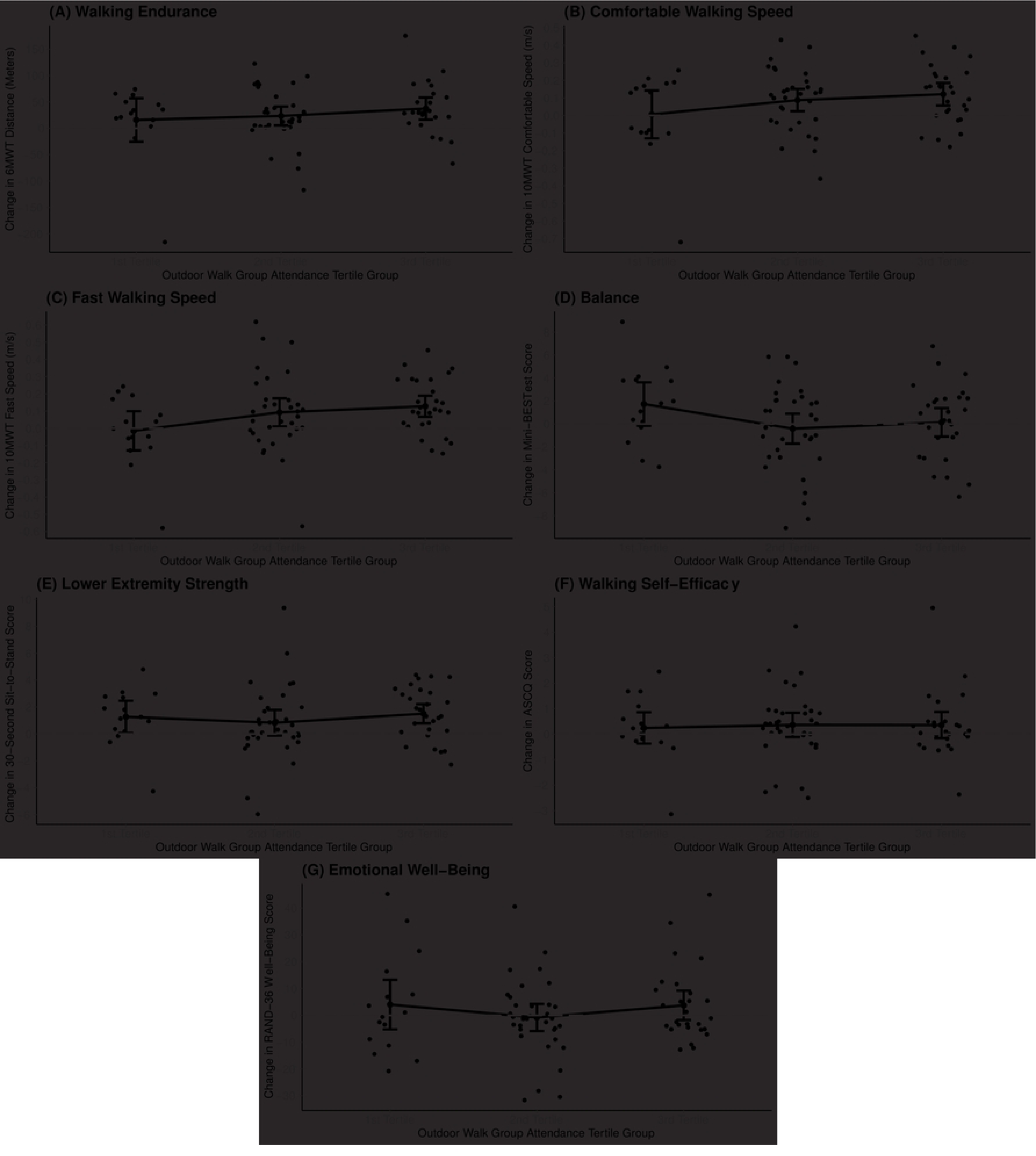
Changes in measures of physical and mental health from baseline to 3 months across three outdoor walk group attendance groups. *Note*: The 1^st^ tertile group attended 0–9 sessions; the 2^nd^ tertile group attended 10–15 sessions; the 3^rd^ tertile group attended 16–20 sessions. Dots above the horizontal dashed line indicate an improvement (change score>0) in the respective health outcome measure from baseline to 3 months.

### Influence of Outdoor Walking Attendance on the Odds of Improvement in Health

Table 3 presents the associations between outdoor walking attendance and the odds of experiencing an improvement in health. Results from the adjusted logistic regression models showed that compared to those attending only 0–9 OWG sessions, the odds of experiencing an improvement in comfortable walking speed from baseline to 3 months tended to be about 6 times larger among participants who attended 16–20 outdoor walking sessions (Odds ratio [OR]=5.97, 95% CI: 1.02, 42.69, *p*=0.06). The odds of experiencing an improvement in fast walking speed were 9 times larger among participants who attended 16–20 outdoor walking sessions than those attending 0–9 sessions (OR=9.00, 95% CI: 1.59, 61.73, *p*=0.02). Likewise, the odds of experiencing an improvement in walking self-efficacy, as measured by the ambulatory self-confidence questionnaire, was about 4 times larger among participants attending 10–15 outdoor walking sessions in comparison to those attending 0–9 sessions (OR=4.03, 95% CI: 1.05, 16.85, *p*=0.047). The raw percentages of participants experiencing an improvement in health across the three attendance tertile groups are presented in Supplementary Fig 1.

**Table 3.** Associations between outdoor walk group attendance and the odds of experiencing improvement in health outcome measures from baseline to 3 months.

| Health outcome measures | Comparisons between OWG attendance tertiles |  |  |
| --- | --- | --- | --- |
|  | 2<sup>nd</sup> tertile (10–15 sessions) | 3<sup>rd</sup> tertile (16–20 sessions) | 3<sup>rd</sup> tertile (16–20 sessions) |
|  | vs | vs | vs |
|  | 1<sup>st</sup> tertile (0–9 sessions) | 1<sup>st</sup> tertile (0–9 sessions) | 2<sup>nd</sup> tertile (10–15 sessions) |
|  | Odds ratio [95% confidence intervals] |  |  |
| 6-minute walk test (m) | 1.92 [0.21, 14.84] | 4.02 [0.36, 45.31] | 2.09 [0.41, 10.62] |
| 10-meter walk test at comfortable pace (m/s) | 3.45 [0.74, 18.56] | 5.97 [1.02, 42.69] † | 1.73 [0.48, 6.23] |
| 10-meter walk test at fast pace (m/s) | 3.10 [0.75, 13.62] | 9.00 [1.59, 61.73] * | 2.91 [0.75, 11.30] |
| Mini-BESTest (0–28) | 0.64 [0.16, 2.58] | 0.94 [0.19, 4.63] | 1.45 [0.47, 4.47] |
| 30-second sit-to-stand (# stands) | 0.39 [0.08, 1.56] | 0.90 [0.15, 4.87] | 2.34 [0.73, 7.48] |
| ASCQ (0–10) | 4.03 [1.05, 16.85] * | 3.08 [0.65, 16.07] | 0.76 [0.23, 2.56] |
| RAND-36 emotional well-being (0–100) | 0.43 [0.11, 1.58] | 0.54 [0.11, 2.45] | 1.26 [0.40, 3.97] |

*Note:* The odds ratios are in reference to the lower tertile/attendance group, with values greater than 1 suggesting potential dose-response relationships. All regression models were adjusted for participant sex, study site, and the Heckman correction. † $p < 0.10$ ; \* $p < 0.05$

### Influence of Outdoor Walking Attendance on the Number of Improvements in Health

Table 4 presents results from the Poisson regression model of the relationship between OWG program attendance and the overall count of improved health from baseline to 3 months. The total number of improvements in the seven health outcomes increased with the number of outdoor walking sessions attended: 3.71±1.61 for 0–9 sessions, 4.03±1.66 for 10–15 sessions, and 4.43±1.50 for 16–20 sessions; the differences across groups, however, were not statistically significant. Analyses conducted with and without the Heckman correction yielded similar results. Therefore, the findings from the models without the correction are presented in Supplementary Tables 2-4.

**Table 4.** Associations between outdoor walk group attendance and the total count of improvement on health outcome measures from baseline to 3 months.

| Outdoor walk group attendance | Total count of improvement on health outcome measures<br>(range: 0–7) |  |
| --- | --- | --- |
| | Mean ± SD | Incident rate ratio [95% CIs] |
| 1<sup>st</sup> tertile (0–9 sessions) | $3.71 \pm 1.61$ | 1.00 (Reference) |
| 2<sup>nd</sup> tertile (10–15 sessions) | $4.03 \pm 1.66$ | 1.18 [0.86, 1.63] |
| 3<sup>rd</sup> tertile (16–20 sessions) | $4.43 \pm 1.50$ | 1.36 [0.94, 1.96] |
| 2<sup>nd</sup> tertile (10–15 sessions) |  | 1.00 (Reference) |
| 3<sup>rd</sup> tertile (16–20 sessions) |  | 1.15 [0.89, 1.50] |

*Note:* CIs = confidence intervals. The incident rate ratios are in reference to the lower tertile/attendance group, with values greater than 1 suggesting potential dose-response relationships. The Poisson regression model was adjusted for participant sex, study site, and the Heckman correction.

## DISCUSSION

This study investigated the potential dose-response relationships between attendance in an OWG program and improvement in seven physical and mental health outcomes among community-dwelling older adults with mobility limitations. Results showed that compared to lower attendance levels, higher attendance was associated with greater improvement in walking endurance and comfortable and fast walking speed before and after the intervention, as well as increased odds of experiencing improvements in comfortable and fast walking speed and walking self-efficacy.

Overall, our findings are in line with the existing body of literature that documents the health benefits of walking interventions [4,42,43] and provides further information about park-based outdoor walk group programs. Firstly, we found that older adults who attended 16–20 outdoor walking sessions exhibited a greater improvement in walking endurance, as measured by the 6-minute walk test, compared to those attending only 0–9 sessions. This finding is similar to those reported by Dondzila et al. [44], where 6-minute walk test performance gradually improved as older adults engaged in higher levels of walking activity per day (e.g., from 2,500 steps/day to 7,500 steps/day). Notably, our study revealed a mean difference of 52.7 meters in 6-minute walk test performance between the high and low attendance groups, which exceeds the established minimal clinically important difference of 30.5 meters for the test [45]. Our study presents preliminary evidence suggesting that participating in 16 or more sessions of a park-based OWG program over a period of 10 weeks may be needed to provide sufficient training for achieving improvements in walking endurance among older adults with mobility limitations. Second, the present study also identified dose-response relationships between the number of outdoor walking sessions attended and improvements in comfortable and fast walking speed in a community setting. While direct comparisons to our findings are limited, several previous studies have explored related concepts [46–49]. For instance, a recent randomized controlled trial led by Klassen and colleagues [47] showed that acute stroke patients receiving high-dose exercise interventions (40 sessions) in an indoor hospital setting exhibited significant improvements in gait speed compared to those receiving standard stroke care, while no such improvements were observed among those with low-dose interventions (20 sessions). Similarly, a meta-analysis involving 2,054 community-dwelling older adults suggested that high-dose therapeutic exercise interventions, defined as three 60-minute sessions per week, had a significant positive impact on improving gait speed, whereas low-dose interventions did not yield similar effects [48]. By highlighting the relationship between attendance in outdoor walking interventions and improvement in physical functioning, our findings provide additional insights into the broader context of exercise dosage and may guide the design, offering, and evaluation of community-based exercise programs aimed to support the health and functional independence of older adults [50,51].

To delve deeper into the impact of attendance in the OWG program, our study expanded its scope beyond merely measuring changes in health. We incorporated two additional approaches to operationalize dose-response and investigate how attendance would affect both the odds and overall count of improvements in health. Through this approach, we identified evidence indicating that attending a greater number of outdoor walking sessions positively influences older adults’ walking self-efficacy, which is a phenomenon not detected using the standard method of measuring absolute change. As a range of methodological approaches have been developed to examine dose-response relationships across fields of health sciences [36–38], further research utilizing distinct operational definitions of response is warranted to evaluate the potential benefits of outdoor walking interventions on health of older adults.

Contrary to expectations, our analysis revealed no significant association between attendance in the OWG program and improvement in balance, lower extremity strength, and emotional well-being. Several factors may shed light on these findings. First and foremost, the OWG program focused primarily on task-specific training designed to enhance older adults’ competency and skills in outdoor ambulation, including walking, turning, and stepping sideways [15]. The targeted nature of our intervention may explain why significant improvements were observed in walking endurance, walking speed, and walking self-efficacy, but not in other general health outcomes. Second, a meta-analysis involving 1,252 participants across 10 studies demonstrated a U-shaped relationship between nature-based exercise and mental health [52]. Particularly, they found that the largest benefits of nature and green exercise were observed at the shortest duration of 5 minutes per day. These benefits diminished as the duration increased to 10-60 min and half-day sessions but rose again for the whole day duration. This phenomenon may partly explain the lack of clear dose-response relationship in our study. Interestingly, an experimental study conducted by Li and colleagues showed contrasting effects of outdoor walking based on timing and location [53]. They found that while walking in an urban environment during nighttime was associated with positive effects on blood pressure, emotions and moods among middle-aged and older adults, walking in the same environment during daytime instead had negative effects on health potentially due to urban stressors such as noise, crowding, and air pollution. As our OWG program was administered in parks within four large Canadian cities during the day, it is possible that the timing and location of the walks may have influenced the observed outcomes. Taken together, these insights highlight the complexity of designing and implementing outdoor walking interventions for older adults, suggesting the need to leverage an integrated paradigm that accounts for the diverse individual, environmental, and contextual factors involved in these initiatives [19].

This study possesses numerous strengths. Our OWG program is among the first that incorporates task-specific outdoor activities targeted towards enhancing community mobility skills among older adults [8]. In addition, our study spanned four major cities across Canada, which strengthens the external validity of our findings and allows generalization to similar urban environments. Moreover, the inclusion of a broad range of outcome measures and operational definitions of response contributes to a better understanding of the impact of outdoor walking interventions on older adults’ health. However, findings should be interpreted within the context of their limitations. For example, although we observed gradients in the association between outdoor walking attendance and total count of improved health outcomes, the lack of significance may result from limited sample size compared to prior studies involving outdoor community ambulation interventions among older adults [8]. Our study was also not designed to establish the optimal attendance level or other dose parameters, such as intensity, frequency, and duration, for outdoor walking interventions. Future research is needed to investigate the impact of attendance in tandem with these dose parameters on the effectiveness of outdoor walking interventions.

In conclusion, this study demonstrates the positive impact of higher attendance in an outdoor walking program on improving walking endurance, walking speed, and walking self-efficacy among older adults with mobility limitations. Our findings reinforce the importance of ongoing clinical and research endeavors to explore factors that may influence outdoor walking attendance and engagement among older adults [11–13,54]. Meanwhile, attention should also be paid to verify how attendance in the intervention programs would influence long-term behavior change. Ultimately, these efforts will allow us to translate volume into value, maximizing the potential of outdoor walking in improving the lives of older adults.

## Data Availability

We did not obtain permission from the Research Ethics Boards at the University of Alberta, University of Manitoba, University of Toronto or McGill University for public availability of the data, nor did we obtain consent for public access to the data from participants. We are therefore unable make the data public. However, data are available on reasonable request and subject to research ethics board review by contacting the corresponding author or the Health Sciences Research Ethics Board at the University of Toronto at.

